# “A cross-sectional case-control study of sexual dysfunction and associated depression in postmenopausal incontinent women.”

**DOI:** 10.1101/2020.07.16.20155564

**Authors:** Tanudeep Kaur, Rajesh Kumari, Jai Bhagwan Sharma, Kavita Pandey, Bharti Uppal, Deb Koushik Sinha, Kumari Anukriti

**Affiliations:** Obstetrics and Gynaecology, AIIMS, New Delhi, India; Psychiatry, AIIMS, New Delhi, India

**Keywords:** Female sexual dysfunction (FSD), urinary incontinence (UI), coital incontinence (CI), depression, sexual health

## Abstract

**Background:** Female sexual dysfunction (FSD) has higher prevalence in postmenopausal women especially with urinary incontinence (UI) and coital incontinence (CI). This study was attempted as there is dearth of literature to quantify FSD, CI and depression in UI women and their correlation with each other especially in Indian scenario.

**Aim:** Aim of this study was to determine the prevalence of FSD, CI and depression in postmenopausal UI women.

**Material and Methods:** Cross-sectional case-control study of 100 menopausal women with previously diagnosed UI with matching controls over period of 3 months were analyzed for the prevalence of FSD, CI and depression using validated questionnaires like Female Sexual Function Index (FSFI) and Primary Health Questionnaire-9 (PHQ-9). Statistical Analysis was performed using SPSS version 19.0.

**Results:** UI is independently associated with FSD, CI and depression (p < 0.001) with prevalence of 100%, 83%, and 100% respectively for cases versus 5%, 6%, and 4% respectively for controls. FSD and CI are also independently associated with depression (p < 0.001). Combined FSD with UI and CI with UI are also significantly associated with depression.

**Conclusion:** UI is independently associated with FSD, CI and depression. FSD and CI are also independently associated with depression. UI is the major determinant of depression in those with both FSD and UI or CI and UI.

**Key Message Points:**

- Female sexual dysfunction is rampant in menopausal women especially the ones who are incontinent, which jeopardizes their quality of life.
- Sexually dysfunctional incontinent females feel socially outcast and are moderate to severely depressed and need addressal.

There is a dearth of literature as regards to this issue in the society which needs to be explored, hence an attempt has been made

## Introduction

Women today live 1/3rd of their lives after menopause. Sexual wellbeing is an important part of postmenopausal woman’s health [1]. Female sexual dysfunction (FSD) as currently defined has a prevalence of 25%-63% amongst all women, with a higher peak in postmenopausal women of 68%-86.5% [2]. Its association with Urinary and Coital incontinence (leaking during intercourse) multiplies it manifold [1,3,4]. FSD is often marred by the four U’s i.e., being unaddressed, underestimated, undiagnosed and untreated [1]. These women are also significantly depressed which also needs addressal and treatment.[5] FSD and depression are amenable to treatment with the treatment of underlying Urinary Incontinence (UI) [3]. There is a dearth of literature regarding quantitative approach to measure FSD in incontinent women and its associated depression especially in Indian scenario. Thus, we aimed to determine the prevalence of FSD, Coital Incontinence (CI) & Depression in postmenopausal urinary incontinent women and their matching continent controls.

## Material and methods

A cross-sectional case-control study was conducted in the urogynecology division of the obstetrics and gynecology department of a tertiary care teaching hospital from 1st February, 2020 to 1st May, 2020 after taking prior permission from the institutional ethics committee (IEC-886/03.01.2020, RP-38/2020) involving 100 previously diagnosed patients of incontinence [Stress UI (SUI)/ Urge UI (UUI)/ Mixed UI (MUI)] as cases and their accompanying relatives without incontinence as matching controls (100) to remove any confounding variables. Taking prevalence rate as 70%, allowable error 10%, sample size required is 171.

The women were enrolled in the study after taking informed written consent on patient information sheet in their understandable language. Patients were enrolled if they had acquired 1) natural menopause and were not on systemic / local hormonal replacement therapy, 2) were willing to talk about their sexual life and depression, 3) had recent heterosexual partnered activity with vaginal intercourse in the past 4 weeks, and 4) had a normal urine routine and culture report.

They were excluded from study if they had 1) a history of pelvic surgery / incontinence surgery / pelvic malignancy / radiation, 2) a history of postmenopausal bleeding, 3) current urinary tract infection / pelvic inflammatory disease/ ≥ stage 2 POPQ prolapse / congenital vaginal abnormalities, 4) sexual dysfunction due to male partner issues, 5) depression due to causes other than UI / FSD, 6) normal bereavement, and 7) history of maniac/ hypo maniac episode or senile dementia.

A detailed history as regards to their demographic profile, marital status, medical and surgical history, gynecological / urological / obstetric / sexual history, menopausal status, and current medications was taken. Detailed general and local examination was done. Permissions were sought beforehand for the use of 2 validated questionnaires – 1) Female Sexual Function Index (FSFI) - the short form of Urogenital Distress Inventory (UDI-6) with 19 questions for subjective validated measure of female sexual function having 6 domains like desire, arousal, lubrication, orgasm, global sexual satisfaction and pain for quantifying FSD [6]; and 2) the Prime-MD Patient Health Questionnaire, 9-item (PHQ-9) for quantifying depression - in conformity with the Diagnostic and Statistical Manual of Mental Disorders, 4th Edition (DSM-IV) especially validated for use in obstetrics and gynecology outpatient clinics [7]. FSFI score ranges from 2 to 36 with a validated cut-off score of </=26.55 signifying Female Sexual Dysfunction (FSD). The PHQ-9 scores of 0-4, 5-9, 10-14, 15-19 and 20-27 represents none or minimal, mild, moderate, moderately severe and severe depression, respectively with a PHQ-9 score of >/= 10 having 88% sensitivity and specificity for major depression. Subjects were also interrogated for Coital incontinence (CI), i.e. leaking during intercourse. Both the questionnaires were not validated in Hindi for use. The permission for their use and verbal translation in Hindi was sought beforehand. The principal investigator translated them verbally in Hindi for patient understanding. Informed and written consent was taken beforehand. The data so obtained was kept confidential, and the whole study was conducted in accordance with the ethical guidelines of Declaration of Helsinki and its amendments. The statistical analysis was performed using SPSS version 19.0. Pearson Chi Square test was applied to assess the various correlations in the data.

## Patient and Public Statement

The development of research question and outcome measures were influenced by patient’s priority, experience and preferences as on interrogation it was evident that sexual dysfunction in these incontinent women bothers them to the core and they wanted to get treated for the same as it takes a toll on their mental health as well and disturbs their quality of life. The appropriate solution of recognition of this issue in these women would help in its resolution. The (n=100) incontinent patients already coming to our urogynaecology OPD were taken as cases and their accompanying relatives by matching were taken as controls after application of appropriate exclusion criteria. The results of our study will benefit patients and study participants in such a way that their often unsaid and unaddressed issue will be addressed and hence appropriate management instituted. Ours was an observational cross-sectional case control study and patients had no burden of intervention.

## Results

Most of our cases were 51-60 years (65%) of age, with all of them being menopausal. The majority of them were married (84%), having a parity of 3 (76%), belonging to low socio-economic status (SES) (62%), with only primary level of education (76%) and having a BMI (Body Mass Index) between 25-29.9 Kg/m2 (70%). Most of them had vaginal route of delivery (91%). The mean duration of incontinence was 17.44 months (range 6 – 60 months). (Table 1).

**Table 1:** Demographic data

| Demographic variable | Cases | Controls |
| --- | --- | --- |
| 1. Age Group (years) |  |  |
| 51-60 | 65 (65%) | 65 (65%) |
| 61-70 | 34 (34%) | 34 (34%) |
| >70 | 1 (1%) | 1 (1%) |
| 2. Marital Status |  |  |
| Married | 84 (84%) | 84 (84%) |
| Never married | 4 (4%) | 4 (4%) |
| Divorced/Separated/ Widowed | 12 (12%) | 12 (12%) |
| 3. Parity |  |  |
| 1 | 3 (3%) | 3 (3%) |
| 2 | 5 (5%) | 5 (5%) |
| 3 | 76 (76%) | 76 (76%) |
| 4 | 10 (10%) | 10 (10%) |
| 5 | 4 (4%) | 4 (4%) |
| 6 | 1 (1%) | 1 (1%) |
| 7 | 1 (1%) | 1 (1%) |
| 4. Socioeconomic status (SES) (Modified Kuppuswamy's SES scale) [8] |  |  |
| Upper | 6 (6%) | 6 (6%) |
| Upper middle | 32 (32%) | 32 (32%) |
| Lower middle | 40 (40%) | 40 (40%) |
| Upper lower | 22 (22%) | 22 (22%) |
| 5. Education |  |  |
| College graduate | 7 (7%) | 7 (7%) |
| Partial college | 17 (17%) | 17 (17%) |
| High school graduate only | 76 (76%) | 76 (76%) |
| 6. Body Mass Index (BMI) (Kg/m<sup>2</sup>) |  |  |
| 18.5-24.9 (Normal weight) | 6 (6%) | 6 (6%) |
| 25-29.9 (Over weight) | 70 (70%) | 70 (70%) |
| 30-34.9 (Obese Type I) | 24 (24%) | 24 (24%) |
| 7. Mode of delivery |  |  |
| Vaginal | 91 (91%) | 91 (91%) |
| Caesarean section | 9 (9%) | 9 (9%) |

Urge urinary incontinence was present in 88% of our cases, and 6% each had SUI and MUI. All (100%) of our UI women (cases) had FSD (FSFI score of </=26.55), whereas in controls only 5% had FSD with 95% having scores of >26.55 (p<0.001, highly significant) (Table 2). Of the various domains of FSFI Scoring, 41% had desire disorders, 16% had arousal disorders, 7% had lubrication issues, 7% had orgasmic issues, 23% complained of a lack of global satisfaction and 6% reported sexual pain issues. Amongst our UI women 83% leaked during sexual intercourse (coital incontinence, CI) compared to only 6% of controls (p<0.001; highly significant) (Table 2). All (100%) of our UI women had depression (PHQ-9 scoring) with as high as 48% having severe depression, and 45% having moderately severe depression versus 0% and only 4% respectively for controls (p<0.001, highly significant) (Table 2).

**Table 2:** Female Sexual Dysfunction (FSD), Coital incontinence (CI) and Primary Health Questionnaire-9 (PHQ-9) scoring in Cases and Controls

| **A. Female Sexual Dysfunction (FSD) in Cases and Controls** |  |  |  |  |  |
| --- | --- | --- | --- | --- | --- |
| FSD | Cases |  | Controls |  | Total |
|  | N | % | N | % |  |
| Present | 100 | 100.0 | 5 | 5.0 | 105 |
| Absent | 0 | 0.0 | 95 | 95.0 | 95 |
| Total | 100 | 100.0 | 100 | 100.0 | 200 |
| $\chi^2 = 180.952$; $df = 1$; $p < 0.001$; Highly significant | | | | | |
| **B. Coital incontinence (CI) in Cases and Controls** |  |  |  |  |  |
| CI | Cases |  | Controls |  | Total |
|  | N | % | N | % |  |
| Present | 83 | 83.0 | 6 | 6.0 | 89 (44.5%) |
| Absent | 17 | 17.0 | 94 | 94.0 | 111 (55.5%) |
| Total | 100 | 100.0 | 100 | 100.0 | 200 |
| $\chi^2 = 101.282$; $df = 1$; $p < 0.001$; Highly significant | | | | | |
| **C. Primary Health Questionnaire-9 (PHQ-9) Scoring for Depression in Cases and Controls** |  |  |  |  |  |
| Depression (PHQ-9) | Cases |  | Controls |  | Total |
|  | N | % | N | % |  |
| None/ Minimal | - | - | 71 | 71.0 | 71 |
| Mild | 2 | 2.0 | 13 | 13.0 | 15 |
| Moderate | 5 | 5.0 | 12 | 12.0 | 17 |
| Moderately severe | 45 | 45.0 | 4 | 4.0 | 49 |
| Severe | 48 | 48.0 | - | - | 48 |
| Total | 100 | 100.0 | 100 | 100.0 | 200 |
| $\chi^2 = 164.255$; $df = 4$; $p < 0.001$; Highly significant | | | | | |
| The PHQ-9 scores in cases varied between 19.41 to 21.17 with a standard error (SE) of 0.442 |  |  |  |  |  |
| The PHQ-9 scores in controls varied between 4.29 to 5.85 with a standard error (SE) of 0.395 |  |  |  |  |  |

We tried to correlate FSD in subjects (cases and controls) with depression (PHQ-9 scoring) and found that in the absence of FSD as high as 70.53% were not depressed, 25.26% were mildly or moderately depressed, only 4.2% were moderately severe depressed and none was severely depressed.

However, in the presence of FSD, severe or moderately severe depression was present in 93.26% (p < 0.001; highly significant) (Table 3). Amongst UI cases, all (100%) had FSD and all of them (100%) were depressed with 48% having severe depression, and 45% having moderately severe depression. Whereas, in those without UI (controls) only 5 patients (20%) had FSD, none of whom had depression except one patient having moderate depression and a non-significant association was found between FSD and depression in them (Table 3).

**Table 3:**
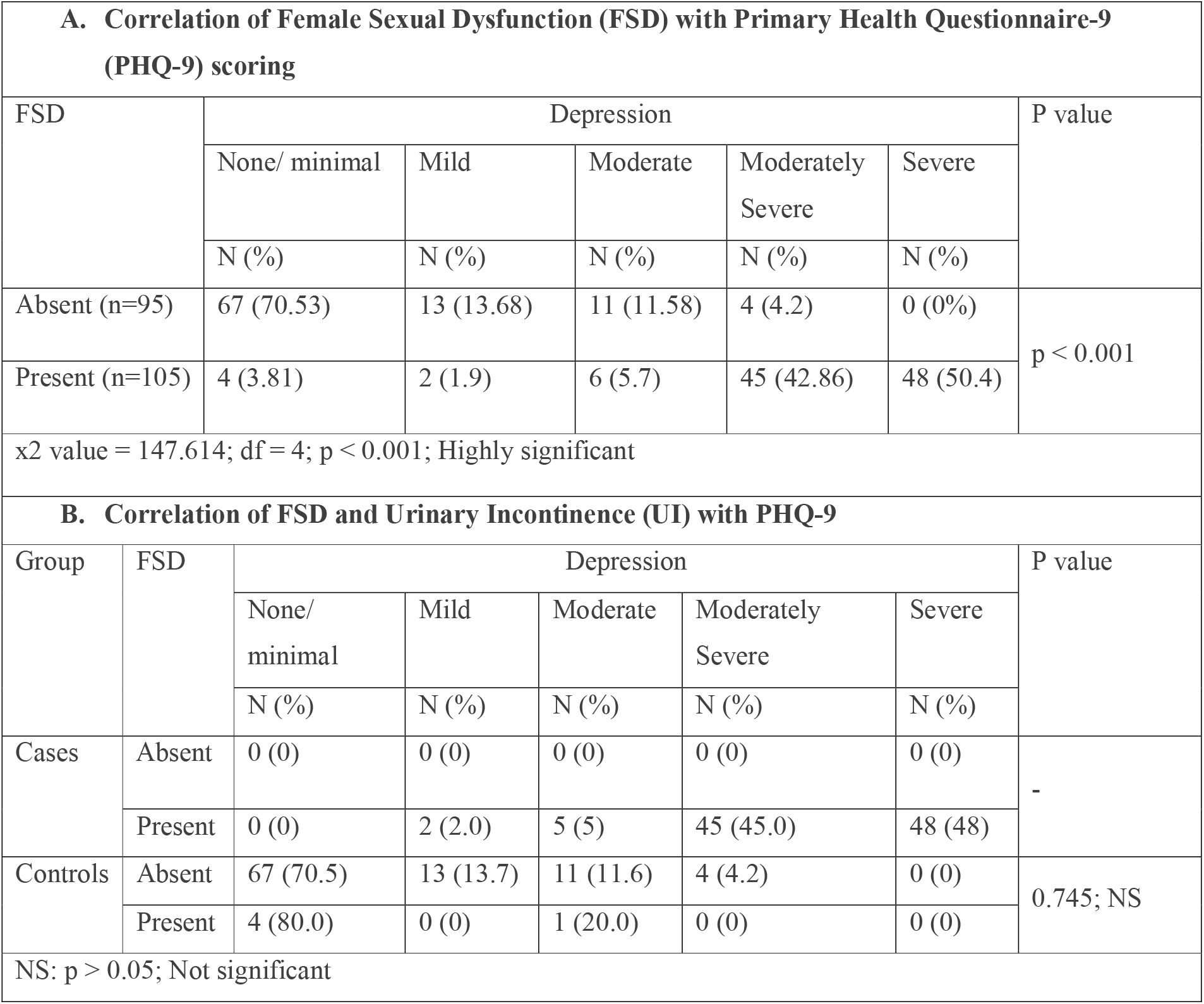
Correlation of Female Sexual Dysfunction (FSD) with Primary Health Questionnaire-9 (PHQ-9) scoring and of FSD and Urinary incontinence (UI) with PHQ-9

Similarly, we tried to correlate CI in subjects (cases and controls) with depression and found that in the absence of CI, 58.56% were not depressed, 23.42% were mildly or moderately depressed, 16.21% were moderately severe depressed and only 1.8% were severely depressed. However, in the presence of CI, severe or moderately severe depression was present in 86.52% (p < 0.001; highly significant) (Table 4).

**Table 4:**
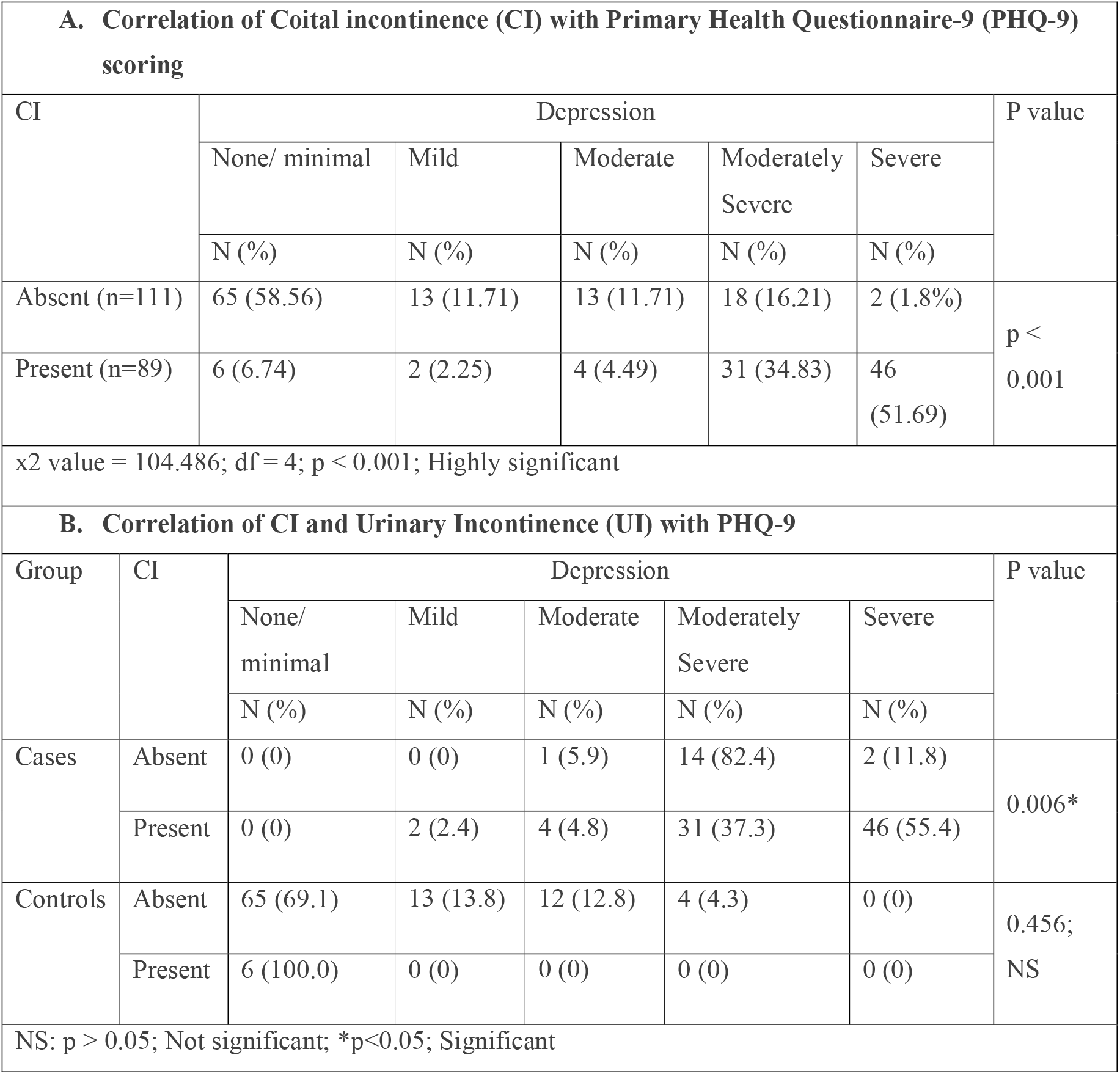
Correlation of Coital incontinence (CI) with Primary Health Questionnaire-9 (PHQ-9) scoring and of CI and Urinary incontinence (UI) with PHQ-9

Amongst UI cases, 83% had CI of whom all had depression with 46% having severe depression and 31% having moderately severe depression and a significant association was found between UI and CI with depression in them (p=0.006, significant). Whereas, in those without UI (controls), 6% had CI, none of whom had depression and a non-significant association was found between CI and depression in them (p=0.456) (Table 4).

## Discussion

Immense literature is available addressing UI in women, but little is known about its impact on sexuality [9], as voicing sexuality is a taboo in Indian society. Though 96% of our sexually dysfunctional incontinent women showed interest in seeking medical advice for their sexual issues, but their primary concern to attend our clinic was still UI. Demographic factors like age, BMI, SES, education, vaginal delivery, self-image perception, and chronic diseases can have a predictive influence on the prevalence of sexual dysfunction, however, similar to other studies we took appropriately matched controls to remove these confounding variables [10]. All (100%) of our UI cases had FSD when quantified on FSFI scoring, whereas only 5% of continent controls had FSD. Our results showed highly significant association between UI and FSD. Literature is abounding with significant association between UI and sexual dissatisfaction, though quantified on different scales. Chun JY et al reported 87% prevalence of sexually dissatisfaction in menopausal women [11]. Temml C et al reported 25.1% prevalence of sexual distress in women of 49.7+/-13.6 years of age, levying a huge socio-economic impact [12]. Laumann EO et al reported sexual dysfunction in 43% of women versus 31% of men [4]. Felippe et al in a similar case-control study as ours of UI cases versus continent controls reported significant association between UI and sexuality when measured on Sexuality Quotient-Female Version Questionnaire (SQ-F) [10]. Amongst FSD, our study showed desire disorders to be the most common (41%), followed by lack of global satisfaction (23%), arousal disorders (16%), lubrication issues (7%), orgasmic issues (7%) and sexual pain issues (6%). Hayes et al in his assessment of 11 studies found desire disorders to be the most prevalent type in 64% (range 16-75%) followed by orgasmic difficulties (35%), arousal disorders (31%) and pain disorders (26%) [13]. Severe UI with its associated decreased libido, vaginal dryness, dyspareunia, decreased satisfaction, orgasmic dysfunction and premature ejaculation are the various reasons cited in literature for the higher prevalence of FSD in UI women [3].

We found significantly higher rates of CI in UI women than continent controls, 83% versus 6% respectively. Serati M et al reported CI prevalence of 10%-27% in his thorough literature search on Pubmed on studies between 1970 to 2008 [14]. He concluded that CI deserves significant attention as it negatively impacts the sexuality. Other studies similarly report the CI prevalence of 56% by Chu CM et al, 24%-66% by Duralde ER et al and 43.7% by Beji et al [3,15,16]. Having UI itself is the greatest predictor of CI [1]. CI can occur during any phase of coitus, but is more commonly seen during penetration in SUI and during orgasm in UUI. CI is amenable to treatment by resolving the underlying UI [14]. Our study could not differentiate between the two due to the recall bias of our patients.

Analysis of depression was a pertinent part of our study. Our UI women had significantly higher rates of depression than continent controls with 100% of our UI women having depression of whom as high as 48% suffered from severe depression and 45% from moderately severe depression versus 0% and only 4% respectively for continent controls. Zorn et al, in his landmark study showed that all (100%) UI patients had significant depression (p<0.001) [17]. Felde et al, in their landmark EPINCONT study also showed higher prevalence of depression in UI women [18]. We also found highly significant association of FSD with depression with 93.26% of our FSD subjects having moderately severe to severe depression, while in the absence of FSD, 70.53% were not depressed (Table 3A). Similar observations have been made by Clayton H, Sreelakshmy K, & Schnatz PF, among many others with statistically significant associations [19,20,21]. When analyzing combined UI and FSD with depression, we found that amongst UI cases, all (100%) had FSD and all (100%) of these sexually dysfunctional incontinent women (i.e., having both FSD and UI) were depressed with 48% having severe depression, and 45% having moderately severe depression. Whereas, in those without UI (controls) only 5 patients had FSD, none of whom had depression except one patient and a non-significant association was found between FSD and depression in them (Table 3B). This may be because the predominant group amongst no FSD group was those without UI (all 95 cases) and the predominant group amongst FSD group was those with UI (100 out of 105 cases). This shows that although FSD is significantly associated with depression (Table 3A); it is even more significant when UI also adds to FSD which causes severe depression in most of the patients having both FSD and UI. In those without UI (controls), the association of FSD with depression did not reach statistical significance because UI is a major factor causing depression, the absence of which makes the association of FSD with depression did not reach its statistical significance. This also shows that underlying UI is the major driving force for both FSD and depression; therefore, diagnosing and treating underlying UI is paramount to the treatment of FSD and depression. Not much literature is published addressing this issue. A closely similar study was done by Cayan S et al who concluded that in UI women there were significantly higher rates of menopausal age group, FSD and depression [22]. We also found highly significant association of CI with depression with 86.52% of CI subjects having moderately severe to severe depression, while in the absence of CI, 58.56% were not depressed (Table 4A).

When analyzing combined UI and CI with depression we found that amongst UI cases, 83% had CI, all of whom had depression with 77% having severe or moderately severe depression and a significant association was found between combined UI and CI with depression. Whereas, in those without UI (controls), 6% had CI with none having depression and a non-significant association was found between CI and depression (Table 4B). The reason for this may be similar to that of combined FSD and UI with depression with the predominant group amongst no CI group being those without UI (94 of 117) and in those amongst CI group being those with UI (83 of 89). Therefore, although CI is significantly associated with depression (Table 4A), it is even more significant when UI also adds to CI. In those without UI (controls), the association of CI with depression did not reach statistical significance because UI is a major factor causing depression, the absence of which makes the association of CI with depression not reach statistical significance. This also shows that similar to FSD, the underlying UI is major driving force for CI and depression; therefore, diagnosing and treating underlying UI is paramount to the treatment of CI and depression. There was dearth of literature as regards these variables but a study by Seung –June oh et al examining the QOL in CI women taking mental health as one of the aspect reported significantly worse outcomes for CI group [23].

We used validated questionnaires and had excellent compliance of cases and controls in answering them. However, ours was a cross-sectional study design measuring the prevalence and not the incidence or causal relationships between UI and FSD, CI and depression thereby limiting the information about the onset of disease and its natural course. FSD, CI and depression are pertinent other variables that significantly affect women presenting with UI even though their main mode of presentation may still be symptoms related to UI. Particular effort to diagnose the magnitude of underlying FSD, CI and depression with validated questionnaires should be made in all such women as these variables significantly affect the outcome of her underlying UI, mental health and sexual life. Thus, it is the duty of every caregiver to assess a menopausal UI woman for sexual dysfunction and depression in every contact, as it is tremendously improving her quality of life.

## Conclusion

UI is independently associated with FSD, CI and depression. FSD and CI are also independently associated with depression. UI is the major determinant of depression in those with both FSD and UI or CI and UI. Every effort should be made to address FSD, CI and depression in all UI patients.

## Data Availability

All data relevant to the study are included in the article and are available upon re.asonable request. Repository name - Dr Tanudeep Kaur.

## Acknowledgement

I want to thank Dr Ravinder Pal Singh for his significant contribution in drafting and proofreading the article.

